# CT-Guided Direct Current Cardioversion for Atrial Arrhythmias During the COVID-19 Pandemic

**DOI:** 10.1101/2022.02.13.22270911

**Authors:** Mahdi Veillet-Chowdhury, Ghaith Sharaf Dabbagh, Stewart M. Benton, April M. Hill, Jefferson H. Lee, Matthew J. Singleton, Gregory P. Fazio, James E. Harvey, Habib Samady, David Singh, Mohammed Y. Khanji, Saman Nazarian, Francesca Pugliese, Edmond Obeng-Gyimah, Brian T. Schuler, C. Anwar A. Chahal

## Abstract

**Objective:** To assess left atrial appendage (LAA) thrombus detection using a novel cardiac computed tomography (CT) protocol specifically prior to direct current cardioversion (DCCV).

**Background:** Transesophageal echocardiography (TEE) is the gold standard in evaluating the LAA prior to DCCV for atrial fibrillation (AF) or flutter (AFL). The COVID-19 pandemic has restricted availability of TEE and anesthesia support.

**Methods:** Prospective cohort of consecutive patients with uncontrolled AF/AFL referred for DCCV from March 2020 to March 2021 were enrolled. CT evaluation of LAA was performed with an initial arterial and subsequent 30-second delayed acquisition to exclude thrombus prior to DCCV. Primary end points were to assess safety and outcomes.

**Results:** A total of 161 patients were included; mean age 69.8 ± 11.1 years; mean CHA_2_DS_2_-VASc 3.4 ± 1.7; and HAS-BLED 1.8 ± 0.9. Median follow-up 175 (105-267) days with zero cardiac-related deaths, and no episodes of TIA or embolic stroke. There was no statistically significant change in mean glomerular filtration rate (GFR) in the study population between the pre- and post-GFR measurements (73.9 ± 21.0 vs 72.7 ± 20.3; p=0.104). Overall mean total dose length product (DLP) was 1042.8 ± 447.5 mGy*cm. Modifying the CT protocol to a narrower 8-cm axial coverage had a statistically significant decrease in total DLP (from 1130.6 ± 464.1 mGy*cm to 802.1 ± 286.4 mGy*cm; P<0.0001).

**Conclusion:** Cardiac CT is both a safe and feasible alternative imaging to TEE for patients prior to DCCV.

**Perspectives:** *Competency in Medical Knowledge:* Cardiac CT is a powerful tool for the evaluation of left atrial appendage and detection of thrombus prior to direct current cardioversion.

*Translational Outlook:* Our protocol was implemented with the restrictions during COVID-19 in mind, yet CT can be a viable tool beyond the pandemic; future randomized clinical trials can bridge the gap between CT and TEE in the setting of cardioversion and help elucidate its safety profile further.

## 1. INTRODUCTION

Synchronized direct current cardioversion (DCCV) for atrial fibrillation (AF) or atrial flutter (AFL) remains a mainstay therapy for selected patients in whom a rhythm control strategy is pursued (1-3). With AF or AFL of >48 hours or unknown duration and without anticoagulation for the preceding 3 weeks, it is recommended to perform transesophageal echocardiography (TEE) to exclude intracardiac thrombus, prior to cardioversion (3). Studies have shown that the thromboembolic risk after cardioversion is highest in the first 72 hours and that the majority of events occur within 10 days. However, these events can occur as late as 4-weeks due to atrial stunning, and therefore it is imperative to ensure full anticoagulation pre- and post-DCCV (4-6).

Sensitivity of TEE is 93-100% and specificity is 99-100% in excluding left atrial appendage (LAA) thrombus (7,8). Although, TEE is considered the gold standard for LAA thrombus clearance prior to DCCV, one of the major drawbacks of TEE evaluation is adequate visualization of all the LAA lobes due to reverberation artifact from a prominent “coumadin ridge” as well as patient comfort and cooperation. Cardiac computed tomography (CT) with dedicated windowing techniques can compensate for these effects and allow a clear delineation of all the LAA lobes on multiple oblique planes. Recently, a large study of 260 patients that evaluated both cardiac CT and TEE prior to radiofrequency ablation for AF which utilized one standard angiographic phase and three delayed acquisitions at 1-, 3-, and 6-min after contrast, demonstrated a sensitivity and negative predictive value of 100%. The specificity and positive predictive value increased to 100% for both at the 6-minute delay acquisition (9).

Other drawbacks of TEE are the associated risks with anesthesia and esophageal complications, given its invasive nature. Furthermore, with the advent of the Coronavirus Disease 19 (COVID-19) pandemic, there is further associated risk of aerosolization of viral particles as TEEs may provoke coughing or gagging. Additionally, the incidence of AF or AFL has been reported to be 10% in patients hospitalized with COVID-19 and associated with increased mortality with a relative risk of 1.77 (10).

Consequently, identification of additional imaging modalities for left atrial and LAA evaluation pre-DCCV may not only meet an important clinical need but also one that has public health interests related to hospital care and utilization of resources. The Society of Cardiovascular Computed Tomography statement on guidance of cardiac CT during the COVID-19 pandemic, suggested use of cardiac CT for evaluation of left atrial appendage in acute atrial arrhythmia prior to restoration of sinus rhythm (11). One study of 52 patients without the use of a delayed CT imaging acquisition demonstrated that cardiac CT in evaluation of LAA prior to cardioversion had a sensitivity, specificity, positive predictive value and negative predictive value of 100%, 84%, 54% and 100%, respectively (12). Recently, a novel protocol which prospectively evaluated 7 patients with cardiac CT for LAA thrombus prior to electrical cardioversion in the emergency department, had no stroke or thromboembolic events at 30 days (13). To address the urgent clinical need in providing cardioversion for patients with AF and AFL, we implemented a novel protocol for CT-guided cardioversion. Herein, we report the findings of our prospective study, which we believe to be the largest single center consecutive cohort, solely assessing feasibility and outcomes of CT-guided cardioversion.

## 2. METHODS

### 2.1. Study population

This study complies with the Declaration of Helsinki, patients were prospectively enrolled in 4DCT data registry, approved by the Institutional Review Board IRB (WellSpan Health IRB00009830, FWA-00008414). The data will be made available upon reasonable request to the corresponding authors. Between March 2020 and March 2021, consecutive adult (≥18 years) patients with AF or AFL with uncontrolled heart rates, with a glomerular filtration rate (GFR) >30 mL/min/1.73m2 or already on chronic dialysis, who were not on therapeutic anticoagulation or had a break in anticoagulation in the preceding 4-weeks were eligible, as they required clearance prior to DCCV. Exclusion criteria: patients unable to continue with a therapeutic regimen of anticoagulation for at least 30-days post-DCCV due to risks of atrial stunning (n=4; **Figure 1**).

**Figure 1:**
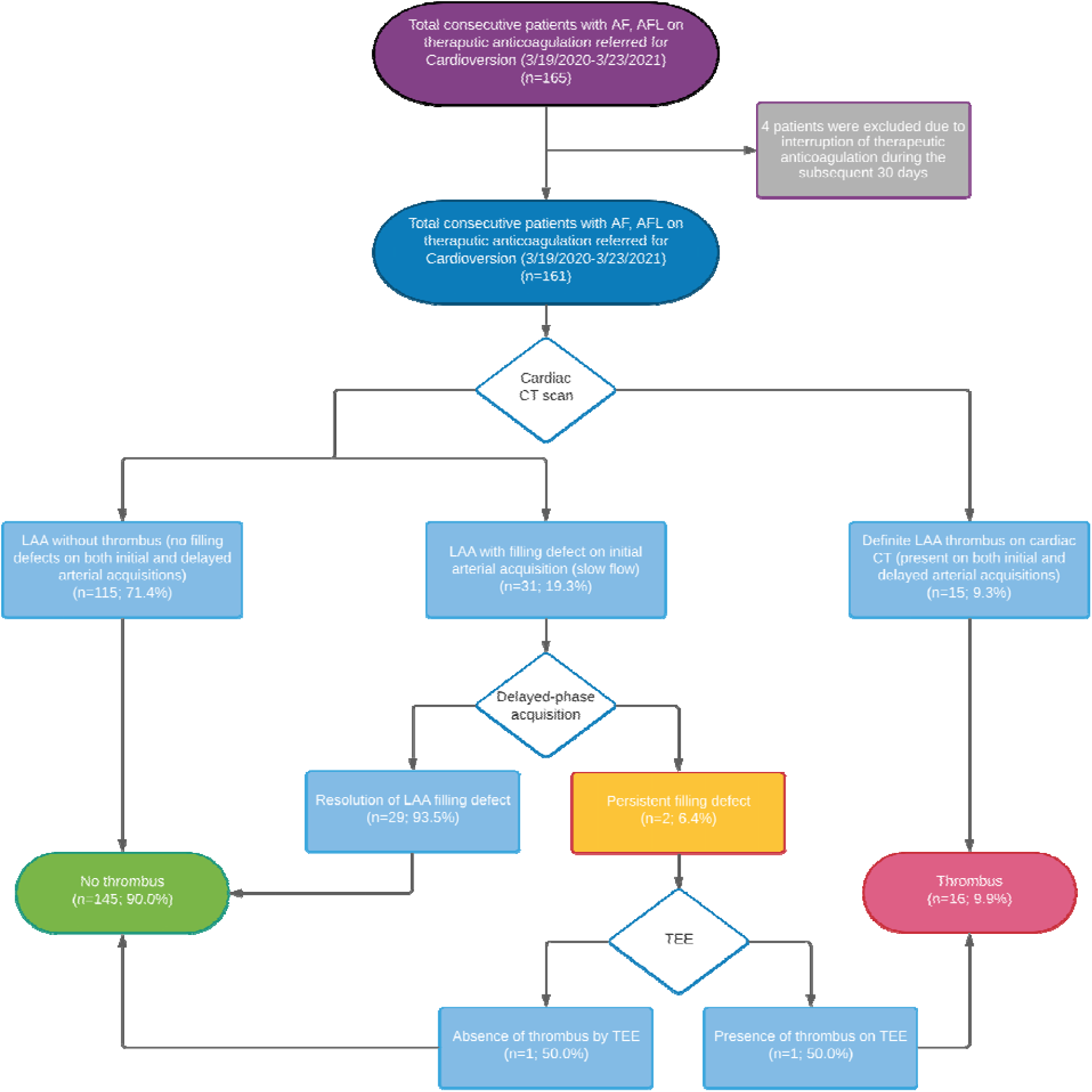
Study Population and Evaluation. (AF: Atrial fibrillation, AFL: Atrial flutter, CT: Computed tomography, LAA: Left atrial appendage, TEE: Transesophageal echocardiography)

### 2.2. CT Protocol

Patients were brought to the novel Alphenix Hybrid Angio/4D-CT (640 slice, Aquilion One™, Canon Medical Systems) suite, and imaged axially using a 320 × 0.5 mm detector configuration, with a gantry rotation time of 275 msec, tube voltage 100-120 kV and auto-tube current maxed at 800 mAs. A contrast-enhanced, prospectively electrocardiogram-triggered exam was performed using 80 milliliter (ml) of iopamidol 370 mg/ml (Isovue-370, Bracco Diagnostics, Princeton, NJ) followed by 80 ml of 0.9% Sodium Chloride injected with a dual injector. Given these patients were in atrial arrhythmias with uncontrolled heart rates at the time of the cardiac CT scan, the acquisition window was adjusted to 40-80% of the R-R interval. The contrast and saline were injected through an 18–20-gauge peripheral intravenous line in the antecubital vein at a rate of 5-6 ml/sec using bolus tracking in the descending aorta at 180 Hounsfield unit (HU) threshold. To help distinguish slow transit of iodinated contrast (“slow flow”) versus thrombus, for all patients, after the initial arterial acquisition was acquired, a 30-second delay was acquired using a smaller diastolic acquisition window. As the goal for initiation of the cardiac CT protocol prior to DCCV was to deliver effective clinical care and minimize resource utilization during the pandemic, the initial cardiac CT protocol applied a 16-cm axial coverage for both the contrast-enhanced acquisition as well as the 30-second delay acquisition to ensure diagnostic quality imaging to avoid TEE use if possible. However, as experience and optimization with the protocol and workflow increased, the 30-second delay acquisition was narrowed to an 8 cm axial coverage of only the LAA on Jan 18, 2021, after 118 patients were completed.

### 2.3. CT analysis protocol

The CT scan was automatically reconstructed using a third-generation iterative reconstruction algorithm, Adaptive Iterative Dose Reduction using 3 Dimensional (AIDR-3D, Canon Medical Systems) in the “standard” setting. Immediately post reconstruction, the CT dataset was analyzed on an imaging post-processing workstation (Vitrea 4.0, Vital Images, Minnetonka, MN, USA) by an advanced imaging cardiologist (COCATS level 3 training [MVC] with 9 years’ experience in cardiac CT). Absence of LAA thrombus was determined by the lack of a visual filling defect within the LAA on the arterial acquisition. In the presence of a filling defect, the 30 second delayed acquisition was analyzed. The filling defect was determined to be “slow flow” if the Hounsfield unit of the filling defect >100 and ≥ the Hounsfield unit of the remainder of the LAA. If the filling defect did not meet these criteria, it was determined to be thrombus or indeterminate (**Figure 2**).

**Figure 2:**
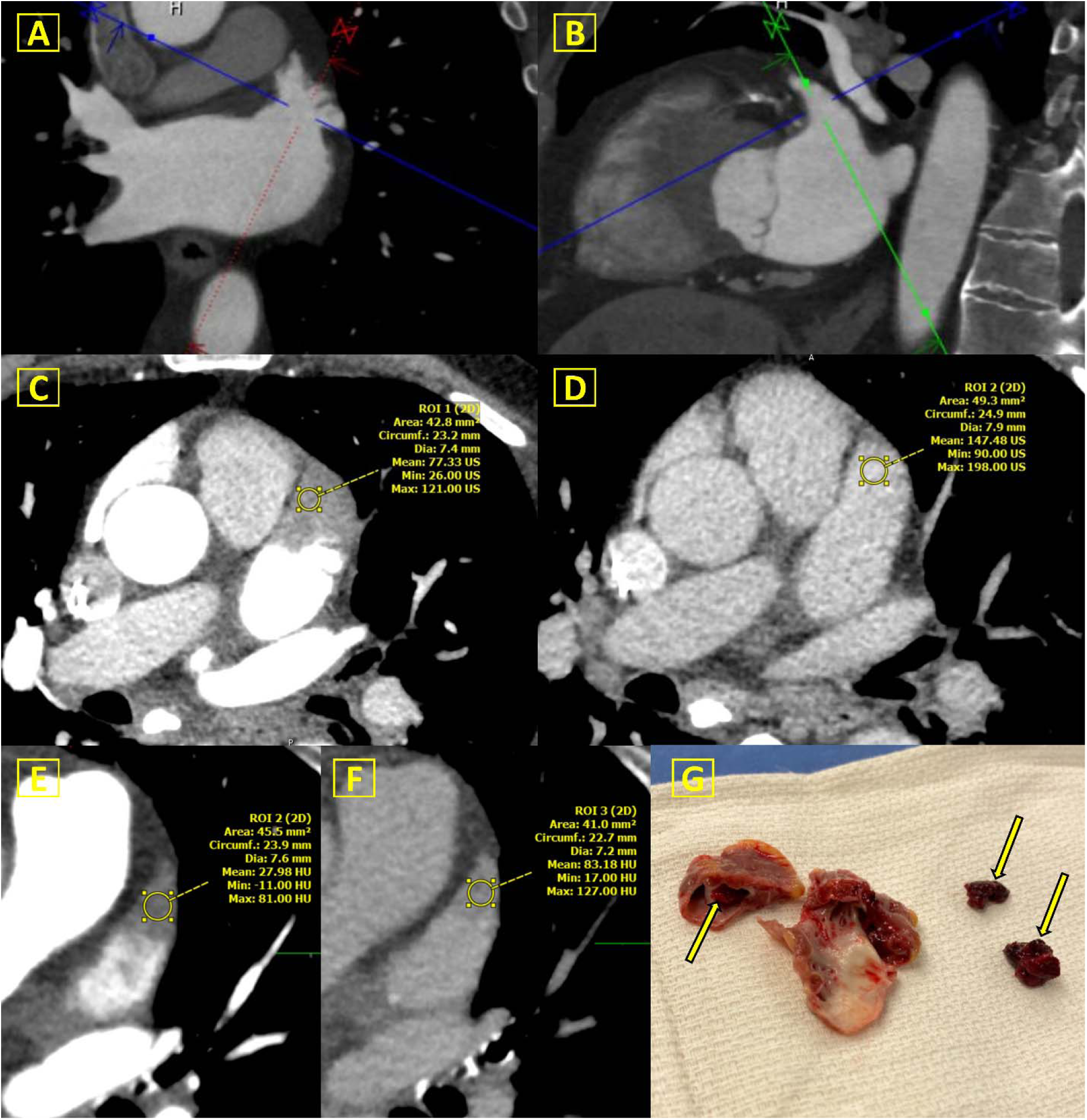
Left Atrial Appendage on Cardiac CT. **Normal CT**: **A**) Initial arterial acquisition cardiac CT demonstrating no evidence of a filling defect or thrombus; **B**) An oblique plane of the multiple lobes of the LAA demonstrating no evidence of filling defect or thrombus **Slow flow**: **C**) Initial arterial acquisition demonstrating a filling defect of the distal LAA lobe with a mean HU of 77; **D**) The 30-second delayed acquisition demonstrating an increase in attenuation with a mean HU of 148, consistent with “slow flow” as opposed to thrombus. **Thrombus: E**) Initial arterial acquisition demonstrating a filling defect of the distal LAA lobe with a mean HU of 28; **F**) The 30-second delayed acquisition demonstrating a persistent low attenuation with a mean HU of 83, consistent with thrombus; **G**) After subsequent 2 cardiac CT scans with similar findings and therapeutic anticoagulation, patient had surgical excision of LAA demonstrating presence of thrombus (**arrows**). (CT: Computed tomography, LAA: Left atrial appendage, HU: Hounsfield units)

A subsequent TEE was performed only as per discretion of the multidisciplinary team that implemented the CT protocol after reviewing data from a relevant study from our center (14). The goals of treatment were focused on providing appropriate clinical care and efficient use of resources with the rapidly spreading COVID-19 pandemic, reducing the need for general anesthesiologists who could focus attention to critical care cases, reducing the risk of aerosolization and endotracheal intubation need for TEE; in contrast CT cardioversion patients were sedated with midazolam and fentanyl by electrophysiologists.

### 2.4. Cardioversion protocol

After the cardiac CT or subsequent TEE confirmed absence of LAA thrombus while on therapeutic anticoagulation, patients received a DCCV in the catheterization suite or adjacent auxiliary procedure room within 2 hours to minimize patient hand-offs and transfers. Patients were then continued on therapeutic anticoagulation for a minimum of 30 days.

### 2.5. Statistical methods

Categorical variables were reported as frequencies and percentages, while continuous variables were reported as means ± standard deviation or median and interquartile range (IQR), depending on distribution. Comparisons between different patient populations were done using Fisher exact or χ2 for categorical variables, and Independent-Samples T test or One-Way ANOVA for continuous variables, as appropriate. When comparing variables between three subgroups, post-hoc analysis was done using residual absolute values over 1.96 to determine significance for categorical variables and the Dunnett method for continuous variables. Survival free of composite stroke and death analysis was conducted using the Kaplan-Meier method and Cox proportional hazard regression, the number of events was small, and the study was underpowered to detect any difference. A two-tailed P value of <0.05 was considered statistically significant. All analyses were performed using IBM SPSS Statistics version 25 (SPSS Inc, IL, USA).

## 3. RESULTS

### 3.1. Patient characteristics

A total of 161 consecutive patients were included in the analysis (mean age 69.8 ± 11.1 years; 101 (62.7%) male). Baseline characteristics and clinical parameters of the study cohort are detailed in **Table** Mean CHA2DS2-VASc and HAS-BLED were, 3.4 ± 1.7 and 1.8 ± 0.9, respectively. There were significant rates of comorbidities, with 81.4% hypertension, 13.0% history of stroke, 17.4% chronic kidney disease, and 31.7% diabetes mellitus. Mean BMI was 31.9 kg/m2 ± 8.0 and the mean left ventricular ejection fraction (LVEF) was 48.6% ± 11.9. Breakdown of the underlying arrhythmia demonstrated 91.9% and with AF and 8.1% with AFL.

### 3.2. Cardiac CT

The cardiac CT evaluation of the LAA is detailed in **Table 2**. The average heart rate during the initial and delayed acquisitions were 97.0 ± 31.0 beats per minute (bpm) and 101.3 ± 29.0 bpm, respectively. As detailed in **Figure 1**, 115 (71.4%) patients had no LAA filling defects on initial and delayed arterial acquisitions; 31 (19.3%) had a filling defect on the initial acquisition “slow flow” that subsequently resolved in 29 (93.5%) on the delayed acquisition and remained persistent in 2 (6.4%). These 2 equivocal cases had a TEE demonstrating thrombus in one case. Definite thrombus on cardiac CT was present on 15 (9.3%) patients based on persistent filling defect with a demarcated border on both initial and delayed acquisitions (**Figure 2e-g**).

**Table 1:** Demographics and Clinical Parameters.

|  |  |
| --- | --- |
| N | 161 |
| Age at scan (y) | 69.8 ± 11.1 |
| Male (%) | 101 (62.7) |
| Hypertension (%) | 131 (81.4) |
| Diabetes Mellitus type 2 (%) | 51 (31.7) |
| Dyslipidemia (%) | 102 (63.4) |
| Prior Stroke or TIA (%) | 21 (13.0) |
| Carotid disease (%) | 2 (1.2) |
| Peripheral Vascular Disease (%) | 9 (5.6) |
| Coronary Artery Disease (%) | 52 (32.3) |
| Ejection Fraction (%) | 48.6 ± 11.9 |
| HFrEF (%) | 41 (25.5) |
| HFpEF (%) | 22 (13.7) |
| Non-valvular Atrial Fibrillation/Flutter (%) | 147 (91.3) |
| Paroxysmal Atrial Fibrillation (%) | 90 (55.9) |
| Persistent Atrial Fibrillation (%) | 58 (36.0) |
| Atrial Flutter (%) | 13 (8.1) |
| Chronic Kidney Disease (%) | 28 (17.4) |
| Dialysis (%) | 3 (1.9) |
| Body mass index (±SD) | 31.9 ± 8.0 |
| Creatinine (IQR) | 0.97 (0.79-1.10) |
| INR (IQR) | 1.10 (1.0-1.2) |
| CHA<sub>2</sub>DS<sub>2</sub>-VASc Score (±SD) | 3.4 ± 1.7 |
| HAS-BLED Score (±SD) | 1.8 ± 0.9 |
| Prior LAAO Watchman (%) | 2 (1.2) |
| Anticoagulation |  |
| Warfarin (%) | 12 (7.5) |
| Apixaban (%) | 108 (67.1) |
| Rivaroxaban (%) | 8 (4.9) |
| Dabigatran (%) | 2 (1.2) |
| Heparin (%) | 24 (14.6) |
| Enoxaparin (%) | 6 (3.7) |
| Edoxaban (%) | 1 (0.6) |
| TIA: Transit ischemic attack, HFrEF: Heart failure with reduced ejection fraction, HFpEF: : Heart failure with preserved ejection fraction, LAAO: Left atrial appendage occlusion, SD: Standard deviation, IQR: Interquartile range |  |

**Table 2:** Cardiac CT evaluation of LAA.

|  |  |
| --- | --- |
| N | 161 |
| CT Contrast dose (ml) | 80.7 ± 7.5 |
| Heart rate on initial arterial acquisition (bpm) | 97.0 ± 31.0 |
| Heart rate on delayed acquisition (bpm) | 101.3 ± 29.0 |
| LAA without thrombus (%) | 115 (71.4) |
| LAA with filling defect on initial arterial acquisition (%) | 31 (19.3) |
| Definite LAA thrombus on cardiac CT (%) | 15 (9.3) |
| Reflex TEE (%) | 2 (1.2) |
| Absence of thrombus by TEE (%) | 1 (50.0) |
| Presence of thrombus on TEE (%) | 1 (50.0) |
| CT: Computed tomography, bpm: Beats per minute, LAA: Left atrial appendage, TEE: Transesophageal echography, SD: Standard deviation, IQR: Interquartile range |  |

The CT scan radiation exposure as represented in the total dose length product (DLP) is detailed both in **Figure 3** and **Table 3**. The overall mean total DLP was 1042.8 ± 447.5 mGy*cm. Modifying the CT protocol to a narrower 8-cm axial coverage on the delayed acquisition had a statistically significant decrease in total DLP (From total DLP 1130.6 ± 464.1 mGy*cm, to total DLP 802.1 ± 286.4 mGy*cm; P<0.0001).

**Figure 3:**
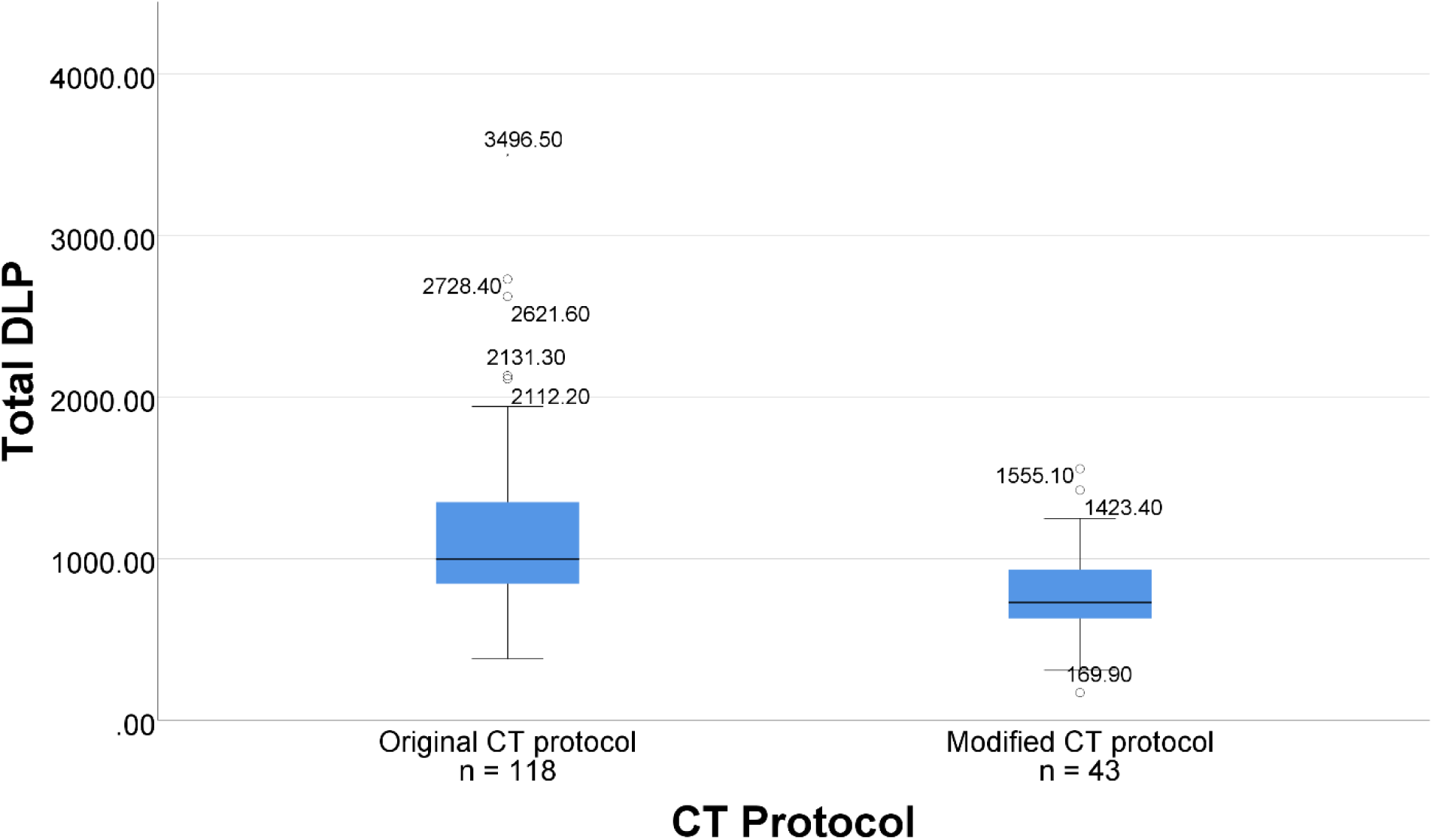
Radiation Dose per CT Protocol. (CT: Computed tomography, DLP: Dose Length Product)

**Table 3:** Radiation doses in total DLP.

|  | **Overall** | **Original CT protocol** | **Modified CT Protocol** | **P-Value** |
| --- | --- | --- | --- | --- |
| **N** | 161 | 118 | 43 |  |
| **Median Total DLP (IQR)** | 933.8 (758.1 - 1222.7) | 995.6 (841.9 – 1350.3) | 727.6 (609.7 – 933.4) |  |
| **Mean Total DLP (±SD)** | 1042.8 ± 447.5 | 1130.6 ± 464.1 | 802.1 ± 286.4 | **<0.0001\*** |
| CT: Computed tomography, DLP: Dose Length Product, SD: Standard deviation, IQR: Interquartile range |  |  |  |  |
| *Obtained through non-parametric test |  |  |  |  |

There were significant differences between the patients depending on the presence of filling defects in the LAA (**Table S1**). The subgroup without any filling defects on both acquisitions had lower rate of prior stroke history compared to both the “slow flow” and definite thrombus subgroups (10 (8.7%) vs. 7 (22.5%) and 4 (26.6%); p=0.032) respectively. This subgroup also had higher LVEF than the other two (51.2 ± 9.6 vs. 44.8 ± 12.2 and 36.4 ± 17.8; p<0.0001), which translated into differences in the rates of HFrEF (22 (19.1%) vs. 11 (35.4%) and 8 (53.3%); p=0.006). Additionally, persistent atrial fibrillation was less prevalent in the first subgroup compared to others (33 (28.6%) vs. 17 (54.8%) and 8 (53.3%); p=0.009) respectively.

### 3.3. Follow up and outcomes

Patient follow-up and outcomes are described in **Table 4**. The patients were followed for a median 175 (105-267) days. Overall, the crude mortality was 7 deaths, of which 0 were cardiac-related. There were no episodes of TIA or embolic stroke during the follow-up period. Univariable and multivariable Cox regression were negative. There was one case of stroke, which occurred at 81 days post-DCCV, despite full anticoagulation, with neurology (stroke) team diagnosis of ischemic stroke due to left carotid atherosclerosis based on CT angiography.

**Table 4:** Follow up and outcomes.

|  |  |
| --- | --- |
| N | 161 |
| Days of follow up (IQR) | 175 (105-267) |
| GFR prior to CT (mL/min/1.73m<sup>2</sup>) | 73.9 ± 21.0 |
| GFR after CT (mL/min/1.73m<sup>2</sup>) | 72.7 ± 20.3 |
| Days between laboratory data (IQR) | 1 (1-15) |
| Acute Renal Failure (%) | 1 (0.6) |
| Stroke/TIA |  |
| Ischemic stroke (%) | 1 (0.6) |
| Embolic stroke (%) | 0 |
| TIA (%) | 0 |
| Non-neurologic embolic event (%) | 0 |
| Death |  |
| Cardiac death (%) | 0 |
| Non-cardiac death (%) | 7 (4.3) |
| GFR: Glomerular filtration rate, CT: Computed tomography, TIA: Transit ischemic attack, SD: Standard deviation, IQR: Interquartile range |  |

There was no statistically significant change in mean GFR in the study population between the pre- and post-CT measurements (73.9 ± 21.0 vs 72.7 ± 20.3; p=0.104). There was one case of acute renal failure in a patient with CKD: the baseline GFR in this patient was 48.6 mL/min/1.73 m2, and at discharge after 19 days was 26.9; the kidney function returned to baseline at the next measurement 65 days post scan and the patient was managed conservatively without the need for renal replacement therapy.

## 4. DISCUSSION

Cardioversion and a rhythm control strategy remain the mainstay therapy for the acute treatment of AF and AFL, necessitating proper evaluation of the LAA. The gold-standard of TEE has been utilized for the past 25 years; however, other non-invasive imaging modalities have revived interest both due to improvement with technology and with the challenges posed by the COVID-19 pandemic. This is the largest single center prospective cohort of consecutive patients undergoing cardiac CT prior to DCCV, with several important findings. First, this approach is safe, feasible and effective in terms of clearance for cardiac thrombus in the acute period. Second, over the long-term (throughout the median follow-up period of 175 days) there were no TIA or embolic strokes and no cardiac-related deaths. Third, the rapid refinement of the scanning protocols and accelerated comparison with TEE, facilitated rapid introduction to a real-world clinical setting. These findings are discussed.

The initial studies that validated the accuracy of TEE in detection of left atrial thrombus were performed in a sample of 231 patients with a thrombi prevalence of 5.2% undergoing elective repair or replacement of the mitral valve or excision of a left atrial tumor, with a sensitivity of 100% and specificity of 99% (8). Another study of 213 patients with a LA/LAA thrombus prevalence of 15%, TEE had a sensitivity of 93.3% and specificity of 100% (7).Therefore, the “gold-standard” of identifying LAA thrombus was initially validated in studies of patients with LAA thrombus of 12 and 30 patients, respectively, non-randomized and observational. The subsequent large study Assessment of Cardioversion Using Transesophageal Echocardiography (ACUTE) was a multicenter, randomized trial of >1,200 patients demonstrating that early TEE-guided cardioversion was safe as compared with the conventional strategy of 4 weeks of anticoagulation (15). In the TEE-guided strategy, there were only two patients with embolic events occurring within the first week after cardioversion, which were associated with AF recurrence and subtherapeutic INRs.

Cardiac CT for LAA clearance has predominantly been evaluated in the context of pre-radiofrequency ablation for AF (16). A meta-analysis of 19 studies with 2955 patients, demonstrated the mean sensitivity, specificity, positive predictive value, and negative predictive value of 96%, 92%, 41% and 99%, respectively. A sub-analysis of 7 studies and 753 patients, which utilized a delayed acquisition imaging protocol, demonstrated the mean sensitivity, specificity, positive predictive value, and negative predictive value 100%, 99%, 92%, and 100%, respectively. Recently, similar to our change in clinical practice due to COVID-19, a large study of 213 patients utilizing cardiac CT prior to AF ablation treatment, the periprocedural cerebrovascular event rate as 0.4% (17).The use of cardiac CT in evaluation of the LAA has also been studied for LAA occlusion (LAAO) device planning. Given the diversity in LAA anatomy, cardiac CT has proven more accurate as compared with TEE in determining device selection (14,18-20).

The major risks associated with a cardiac CT as opposed to a TEE are contrast-induced nephropathy and radiation exposure. In this study, only 1 patient had an episode of acute on chronic renal failure, which returned to baseline on subsequent laboratory evaluation, and without the need for renal replacement therapy. The radiation dose for the cardiac CT was higher as compared to the acceptable doses reported in prior studies, which was due to the larger 16-cm axial acquisition in the initial cohort of patients combined with the fact patients were in rapid AF and AFL during the scan, which requires a wider R-R interval window during acquisition (21,22). This was driven by the clinical need to obtain a definitive diagnostic scan on patients during the ongoing COVID-19 pandemic toward efficient utilization of hospital resources. However, after the protocol was altered with a reduction to an 8-cm axial coverage during the 30-second delay acquisition, the radiation dose was reduced significantly. Although two-dimensional TEE has improved temporal resolution as compared with a gated cardiac CT, more pronounced in patients with elevated heart rates, we were able to obtain diagnostic quality imaging on patients with a mean HR of 97 bpm on the initial arterial acquisition, demonstrating that widening the R-R interval window helped counteract the inherent limitations in temporal resolution of cardiac CT.

An interesting finding in our study is the association between the prior history of stroke and characteristics of the filling defects on the cardiac CT. Similar to LAA blood flow velocity interrogated on TEE with doppler (23), the presence of “slow flow” or definite thrombus on the cardiac CT was associated with prior stroke history. These 2 patient subgroups also had lower LVEF and increased prevalence of persistent atrial fibrillation, which highlights the interplay between LA and LAA function with thromboembolic risk.

With the COVID-19 pandemic suddenly straining hospitals and healthcare resources without significant preparation beginning in March 2020, our system decided to implement cardiac CT driven cardioversion for patients with uncontrolled AF or AFL (24). Given the local and national shortages of PPE, and unclear modes of viral transmission, cardiac CT allowed diversion of essential resources and personnel toward other procedures instead of TEEs. This enabled us to treat patients who could not be managed with rate control strategies or failed anti-arrhythmic medications. While understandably efforts had to be directed to the immediate problem of the global pandemic, patients with other diseases had to be managed. The old proverb of ‘necessity is the mother of invention’ has been epitomized by the pandemic catalyzing the introduction of novel sequences and protocols in healthcare.

### Strengths and limitations

Our study has a number of strengths, including the introduction of a novel CT-guided protocol for DCCV, in a safe and effective manner, and the prospective design. However, there are several limitations including those inherent to an observational design. A major limitation of this study is the lack of a direct correlation with the established “gold standard” of TEE in LAA evaluation. However, as described earlier, due to the resource constraints from the COVID-19 pandemic and associated ethical concerns in performing studies for validation that potentially increase patient and healthcare staff risk, a direct TEE correlation was unable to be performed. Therefore, as the pandemic improves, a larger multi-center study with direct correlation with TEE will be of great benefit and interest for a cost-effective analysis. Additionally, although one risk of cardiac CT is acute renal dysfunction, this study is underpowered to assess this definitively.

## 5. CONCLUSION

A CT-guided DCCV protocol is safe and effective alternative to TEE for the management of patients with atrial tachy-arrhythmias who have failed medical therapy. This protocol was rapidly implemented in the COVID-19 pandemic allowing redirection of resources and staff to COVID-19 cases.

## Data Availability

The data will be made available upon reasonable request to the corresponding authors

## Acknowledgements

none

## Abbreviations

AF: atrial fibrillation
AFL: atrial flutter
CKD: chronic kidney disease
CT: computed tomography
COVID-19: coronavirus disease 19
DCCV: direct current cardioversion
DLP: dose length product
GFR: glomerular filtration rate
HU: hounsfield unit
LAA: left atrial appendage
LVEF: left ventricular ejection fraction
TIA: transient ischemic attack
TEE: transesophageal echocardiography

---

**Table S1:** Comparison of Patient Subgroups with and without LAA filling defects.

|  | **Overall** | **LAA without thrombus** | **LAA with filling defect on initial arterial acquisition (slow flow)** | **Definite LAA thrombus on cardiac CT** | **p-value** |
| --- | --- | --- | --- | --- | --- |
| N (%) | 161 | 115 (71.4) | 31 (19.2) | 15 (9.3) |  |
| Age at scan (y) | 69.8 ± 11.1 | 69.2 ± 11.3 | 72.0 ± 10.7 | 69.7 ± 11.2 | 0.472 |
| Male (%) | 101 (62.7) | 69 (60) | 21 (67.7) | 11 (73.3) | 0.491 |
| Hypertension (%) | 131 (81.4) | 96 (83.4) | 25 (80.6) | 10 (66.6) | 0.288 |
| Diabetes Mellitus type 2 (%) | 51 (31.7) | 39 (33.9) | 6 (19.3) | 6 (40) | 0.232 |
| Dyslipidemia (%) | 102 (63.4) | 76 (66) | 18 (58) | 8 (53.3) | 0.498 |
| Prior Stroke or TIA (%) | 21 (13.0) | **10 (8.7)** | 7 (22.5) | 4 (26.6) | **0.032** |
| Carotid disease (%) | 2 (1.2) | 2 (1.7) | 0 | 0 | 0.667 |
| Peripheral Vascular Disease (%) | 9 (5.6) | 6 (5.2) | 3 (9.6) | 0 | 0.387 |
| Coronary Artery Disease (%) | 52 (32.3) | 36 (31.3) | 10 (32.2) | 6 (40) | 0.795 |
| Ejection Fraction (%) | 48.6 ± 11.9 | **51.2 ± 9.6** | 44.8 ± 12.2 | 36.4 ± 17.8 | **<0.0001** |
| HFrEF (%) | 41 (25.5) | **22 (19.1)** | 11 (35.4) | **8 (53.3)** | **0.006** |
| HFpEF (%) | 22 (13.7) | 15 (13) | 6 (19.3) | 1 (6.6) | 0.470 |
| Non-valvular Atrial Fibrillation/Flutter (%) | 147 (91.3) | 105 (91.3) | 29 (93.5) | 13 (86.6) | 0.740 |
| Paroxysmal Atrial Fibrillation (%) | 90 (55.9) | 71 (61.7) | 13 (41.9) | 6 (40) | 0.061 |
| Persistent Atrial Fibrillation (%) | 58 (36.0) | **33 (28.6)** | **17 (54.8)** | 8 (53.3) | **0.009** |
| Atrial Flutter (%) | 13 (8.1) | 11 (9.5) | 1 (3.2) | 1 (6.6) | 0.505 |
| Chronic Kidney Disease (%) | 28 (17.4) | 19 (16.5) | 5 (16.1) | 4 (26.6) | 0.609 |
| Dialysis (%) | 3 (1.9) | 2 (1.7) | 0 | 1 (6.6) | 0.288 |
| Body mass index | 31.9 ± 8.0 | 32.3 ± 8.0 | 31.1 ± 8.1 | 31.2 ± 8.2 | 0.739 |
| Creatinine | 2.1 ± 9.7 | 1.7 ± 7.6 | 3.9 ± 17.7 | 1.3 ± 1.1 | 0.507 |
| INR | 1.2 ± 0.4 | 1.2 ± 0.3 | 1.2 ± 0.2 | 1.3 ± 0.7 | 0.254 |
| CHA<sub>2</sub>DS<sub>2</sub>-VASc Score | 3.4 ± 1.7 | 3.2 ± 1.6 | 3.6 ± 1.8 | 3.8 ± 1.9 | 0.315 |
| HAS-BLED Score | 1.8 ± 0.9 | 1.7 ± 0.9 | 1.9 ± 0.8 | 2.0 ± 0.6 | 0.473 |
| CT: Computed tomography, LAA: Left atrial appendage, TIA: Transit ischemic attack, HFrEF: Heart failure with reduced ejection fraction, HFpEF: : Heart failure with preserved ejection fraction<br>*Italicized values remained significant in post-hoc analysis between the three groups.* |  |  |  |  |  |

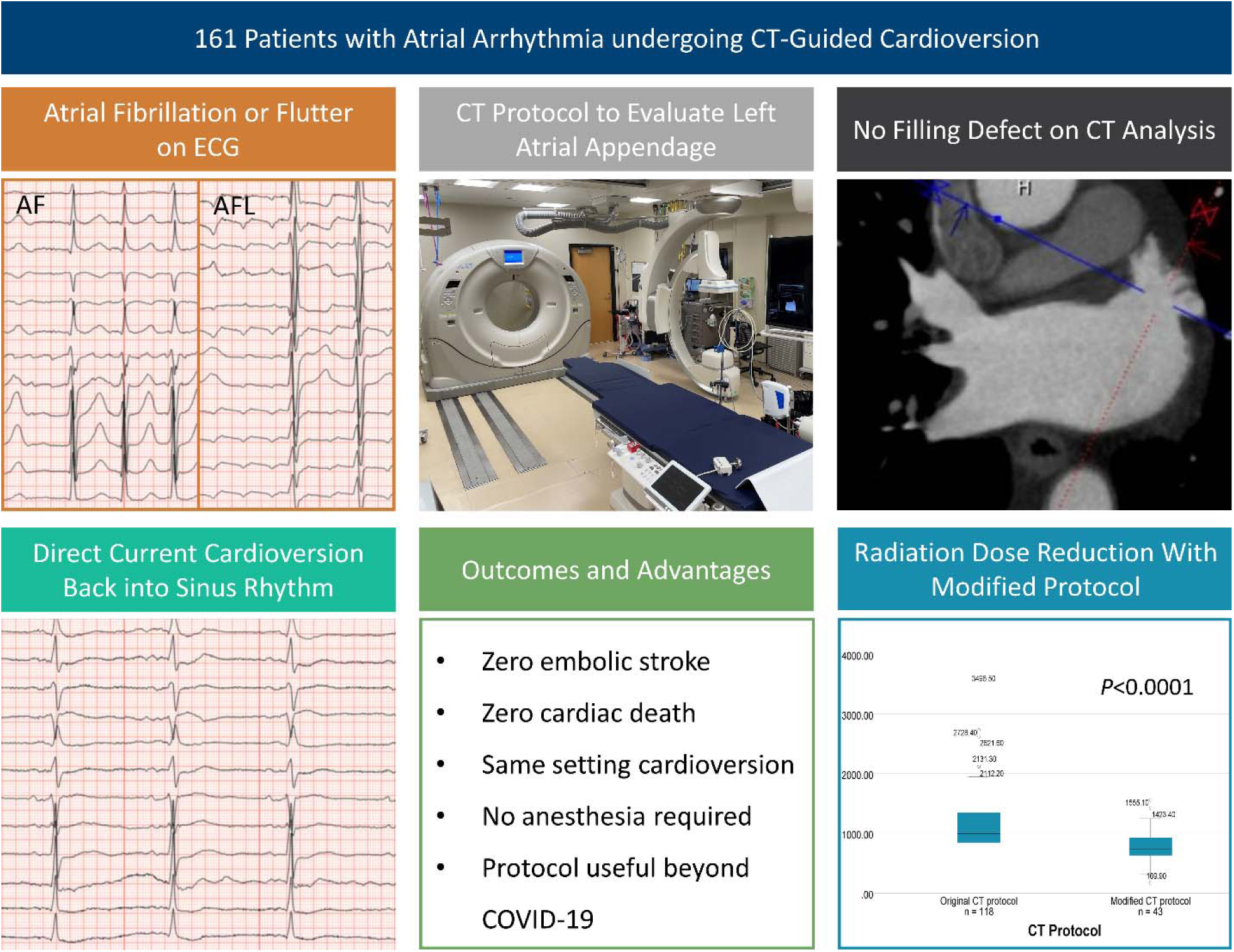

**Central Illustration: CT-Guided Cardioversion Protocol Key Steps and Outcomes**

(CT: Computed tomography, AF: Atrial fibrillation, AFL: Atrial flutter, ECG: Electrocardiogram)

## REFERNCES

1. Hagens VE, Vermeulen KM, TenVergert EM et al. Rate control is more cost-effective than rhythm control for patients with persistent atrial fibrillation--results from the RAte Control versus Electrical cardioversion (RACE) study. Eur Heart J 2004;25:1542–9.

2. Van Gelder IC, Haegeli LM, Brandes A et al. Rationale and current perspective for early rhythm control therapy in atrial fibrillation. Europace 2011;13:1517–25.

3. January CT, Wann LS, Alpert JS et al. 2014 AHA/ACC/HRS guideline for the management of patients with atrial fibrillation: a report of the American College of Cardiology/American Heart Association Task Force on practice guidelines and the Heart Rhythm Society. Circulation 2014;130:e199–267.

4. Klein AL, Grimm RA, Murray RD et al. Use of transesophageal echocardiography to guide cardioversion in patients with atrial fibrillation. N Engl J Med 2001;344:1411–20.

5. Berger M, Schweitzer P. Timing of thromboembolic events after electrical cardioversion of atrial fibrillation or flutter: a retrospective analysis. Am J Cardiol 1998;82:1545–7, A8.

6. Airaksinen KE, Gronberg T, Nuotio I et al. Thromboembolic complications after cardioversion of acute atrial fibrillation: the FinCV (Finnish CardioVersion) study. J Am Coll Cardiol 2013;62:1187–92.

7. Hwang JJ, Chen JJ, Lin SC et al. Diagnostic accuracy of transesophageal echocardiography for detecting left atrial thrombi in patients with rheumatic heart disease having undergone mitral valve operations. Am J Cardiol 1993;72:677–81.

8. Manning WJ, Weintraub RM, Waksmonski CA et al. Accuracy of transesophageal echocardiography for identifying left atrial thrombi. A prospective, intraoperative study. Ann Intern Med 1995;123:817–22.

9. Spagnolo P, Giglio M, Di Marco D et al. Diagnosis of left atrial appendage thrombus in patients with atrial fibrillation: delayed contrast-enhanced cardiac CT. Eur Radiol 2021;31:1236–1244.

10. Musikantow DR, Turagam MK, Sartori S et al. Atrial Fibrillation in Patients Hospitalized With COVID-19: Incidence, Predictors, Outcomes, and Comparison to Influenza. JACC Clin Electrophysiol 2021;7:1120–1130.

11. Choi AD, Abbara S, Branch KR et al. Society of Cardiovascular Computed Tomography guidance for use of cardiac computed tomography amidst the COVID-19 pandemic Endorsed by the American College of Cardiology. J Cardiovasc Comput Tomogr 2020;14:101–104.

12. Achenbach S, Sacher D, Ropers D et al. Electron beam computed tomography for the detection of left atrial thrombi in patients with atrial fibrillation. Heart 2004;90:1477–8.

13. Christians BE, Solie CJ, Swanson MB et al. The Iowa less aggressive protocol: A mixed-methods study on the novel treatment protocol of atrial fibrillation. Am J Emerg Med 2021;45:439–445.

14. Veillet-Chowdhury M, Benton SM, Jr., Chahal CAA et al. Intraprocedural Hybrid Cardiac Computed Tomography for Left Atrial Appendage Occlusion: A Concept and Feasibility Study. JACC Cardiovasc Interv 2021;14:1852–1853.

15. Kleiman NS, Califf RM. Results from late-breaking clinical trials sessions at ACCIS 2000 and ACC 2000. American College of Cardiology. J Am Coll Cardiol 2000;36:310–25.

16. Romero J, Husain SA, Kelesidis I, Sanz J, Medina HM, Garcia MJ. Detection of left atrial appendage thrombus by cardiac computed tomography in patients with atrial fibrillation: a meta-analysis. Circ Cardiovasc Imaging 2013;6:185–94.

17. Akhtar T, Wallace R, Daimee UA et al. Transition from transesophageal echocardiography to cardiac computed tomography for the evaluation of left atrial appendage thrombus prior to atrial fibrillation ablation and incidence of cerebrovascular events during the COVID-19 pandemic. J Cardiovasc Electrophysiol 2021.

18. Eng MH, Wang DD, Greenbaum AB et al. Prospective, randomized comparison of 3-dimensional computed tomography guidance versus TEE data for left atrial appendage occlusion (PRO3DLAAO). Catheter Cardiovasc Interv 2018;92:401–407.

19. Chow DH, Bieliauskas G, Sawaya FJ et al. A comparative study of different imaging modalities for successful percutaneous left atrial appendage closure. Open Heart 2017;4:e000627.

20. Saw J, Fahmy P, Spencer R et al. Comparing Measurements of CT Angiography, TEE, and Fluoroscopy of the Left Atrial Appendage for Percutaneous Closure. J Cardiovasc Electrophysiol 2016;27:414–22.

21. Rybicki FJ, Otero HJ, Steigner ML et al. Initial evaluation of coronary images from 320-detector row computed tomography. Int J Cardiovasc Imaging 2008;24:535–46.

22. Einstein AJ. Effects of radiation exposure from cardiac imaging: how good are the dataã J Am Coll Cardiol 2012;59:553–65.

23. Fatkin D, Kelly RP, Feneley MP. Relations between left atrial appendage blood flow velocity, spontaneous echocardiographic contrast and thromboembolic risk in vivo. J Am Coll Cardiol 1994;23:961–9.

24. Siripanthong B, Hanff TC, Levin MG et al. Coronavirus disease 2019 is delaying the diagnosis and management of chest pain, acute coronary syndromes, myocarditis and heart failure. Future Cardiol 2021;17:3–6.

